# TRAJECTORY AND PREDICTORS OF MATERNAL RELATIONSHIP SATISFACTION FROM PREGNANCY TO FIVE YEARS POSTPARTUM

**DOI:** 10.64898/2026.07.01.26356966

**Authors:** Meagan Emily Crowther, Sienna Cherry OW Starkie, Donna M Pinnington, Sean PA Drummond, Bei Bei

**Affiliations:** School of Psychological Sciences, Faculty of Medicine, Nursing and Health Sciences, Monash University, Clayton, Australia

## Abstract

**Purpose:** Relationship satisfaction is a key factor in women’s mental health and wellbeing outcomes, however, relationship satisfaction may change in times of stress and uncertainty, such as the perinatal period. While positive romantic relationships during the perinatal period may offer benefits, changes in relationship satisfaction and what may contribute to them longitudinally is not currently well understood.

**Method:** One hundred and fifty-seven women in late pregnancy (∼30 weeks gestation) took part in a longitudinal analysis of relationship satisfaction from pregnancy to five years postpartum, across eight timepoints. The study is a secondary analysis of a randomised controlled trial of sleep and diet intervention. Latent growth modelling was used to examine the trajectory of relationship satisfaction change over time and explore predictors of this trajectory.

**Results:** There was a significant decline in relationship satisfaction both during pregnancy and the postpartum. Being of non-White race was associated with overall lower relationship satisfaction (*p* = 0.004). While being less financially comfortable (*p* = 0.007), having less than a postgraduate degree (*p* = 0.049), and older age (*p* = 0.026) were associated with steeper decrease in relationship satisfaction during the postpartum.

**Conclusion:** This current study demonstrates that relationship satisfaction declines from pregnancy to five years postpartum in a sample of Australian mothers. Minority race, financial comfortability, education level and age may influence the magnitude of this trajectory. These findings highlight a need to better support couples during this challenging time, not just through pregnancy and birth, but also through early childhood.

## INTRODUCTION

Romantic relationships are a pivotal part of many adults’ lives. Better relationship satisfaction is associated with better overall health (Robles et al., 2014), well-being (Be et al., 2013), lower risk of early mortality (Robles et al., 2014), and better postpartum health (Tissera et al., 2021). Relationship satisfaction, the measure of how content an individual is, or couple are, with their romantic relationship, changes dynamically over the life of the relationship (Bühler et al., 2021). These changes in satisfaction frequently coincide with periods of stress, experienced either by the individual (e.g., work stress) or by the dyad (e.g., financial hardship). One such life event is the transition to parenthood.

The transition to parenthood can be a challenging time for couples, as both members of the partnership experience significant change and often stress. Becoming a parent is fulfilling and exciting for many (Porat-Zyman et al., 2017), but it also requires the adjustment to new social roles and the demands of infant care (Kluwer, 2010). Welcoming the first child requires both parents’ re-assessment of their self-identity, as well as re-structuring of the family unit and interpersonal relationships (Saxbe et al., 2018). Additionally, during this time some families are faced with additional challenges such as financial stress, birth complications, premature birth and isolation from families, which may exacerbate relationship stress during this period.

Existing research has predominately focused on examining relationship satisfaction during early (one to two years) postpartum, which indicates a decline in relationship satisfaction (Mitnick et al., 2009; Twenge et al., 2003). This period is marked by the challenges of early parenthood such as night infant caregiving, sleep loss (Bei et al., 2015), and shifting of personal roles (Saxbe et al., 2018). There is less research beyond infancy into early childhood, when the demands on individuals and the couple dynamic may change (Bogdan et al., 2022). Cross sectional research shows that as a couple moves beyond the early infancy stage, some couples show a lower relationship satisfaction compared to those parenting infants (Han et al., 2023; Meyer et al., 2016). However, there is a lack of longitudinal research of relationship satisfaction from prenatal to early childhood aged parenting. Therefore, given the protective benefits of relationship satisfaction to an individual’s health and wellbeing, there is a need to examine the trajectory of relationship satisfaction during the transition to parenthood beyond two years postpartum.

Furthermore, there is a need to explore potential predictors of the trajectory of relationship satisfaction during the transition to parenthood. Given that factors such as age, financial strain, being part of a minority group, mental health history, as well as infant and birth related factors, may increase vulnerability during the transition to parenthood (Gopalan et al., 2022; Saxbe et al., 2018), there is a need to explore whether these factors may meaningfully contribute to the trajectory of relationship satisfaction. This study aimed to (1) examine the trajectory of relationship satisfaction from third trimester of pregnancy to five years postpartum, and (2) identify factors which may be associated with the trajectory of relationship satisfaction during the first five years postpartum.

## METHODS

This is a secondary analysis of a Randomised Controlled Trial which explored the effect of a Cognitive Behavioural Therapy for better sleep vs Healthy Diet intervention during the perinatal period. The RCT protocol (Bei et al., 2019), primary (Bei et al., 2023) and other outcomes (Astbury et al., 2022, 2025; Quin et al., 2022; Verma et al., 2024) have been published previously. This study was retrospectively registered (ACTRN12616001462471) on 19/10/2016, after the first participant enrolled on 16/5/2016 but before the end of recruitment on 11/01/2017. The delay in registration was due to limited resources during the early phase of recruitment, when recruitment rate was substantially higher than expected. This study was conducted in line with the Australian National Statement on Ethical Conduct (2007) and ethics approval was attained from the Royal Women’s Hospital and Monash University Human Research Ethics Committees.

### Participants

Participants were a community sample of expectant parents, recruited from Childbirth Education Centre at a Victorian public maternity hospital (Royal Women’s Hospital). Inclusion Criteria were: (a) Nulliparas (i.e., not having given birth to older children); (b) Age 18 or above; (c) Singleton pregnancy; (d) Able to read and write in English; (e) Have regular access to email and internet. Exclusion criteria: (a) The use of medications or substances that directly affect sleep (including sleep medications, melatonin, steroid inhalers, antidepressant medications, cannabis, etc.); (b) Unstable medical conditions that directly affect sleep; (c) Participants who endorse symptoms of, or are known to have significant physiologically based sleep disorders (e.g., sleep apnea, see protocol (Bei et al., 2019) for further details). (d) Participants with fixed night shift work between midnight and 5 am, or rotating work schedules that require night shifts during the study period. (e) Participants with any of the following mental health conditions as determined by structured clinical interview: Major Depressive Disorder (current); Posttraumatic Stress Disorder (current); Panic Disorder if associated with nocturnal panic attacks > 4 times in the past month; Bipolar Disorder (lifetime); Psychotic Disorders (lifetime); Substance Use Disorders (during pregnancy). For the present analysis, only participants who reported being in a relationship (married or de facto) at baseline were included.

### Procedure

All participants provided informed consent online and eligibility was determined via structured telephone interview. All surveys were completed online. Eligible participants were randomised 1:1 to a “Healthy Sleep” (therapist-assisted Cognitive Behavioural Therapy) or “Healthy Diet” (active control) intervention. Both groups received a one-hour telephone orientation with either a psychologist (Healthy Sleep) or dietitian (Healthy Diet), followed by intervention emails delivered at six key perinatal milestones (Bei et al., 2019). Neither of these interventions included content that targeted relationship satisfaction. Repeated assessment of outcomes of interest took place at T1 (30 weeks gestation), T2 (35 weeks gestation), T3 (1.5 months postpartum), T4 (3 months postpartum), T5 (6 months postpartum), T6 (12 months postpartum), T7 (24 months postpartum) and T8 (5 years postpartum).

### Measures

Demographic details were collected at baseline which included: age, level of education, yearly household income, ethnicity, relationship status. Previous mental health history was derived from the *Mini International Neuropsychiatric Interview* (Sheehan et al., 1997). Financial comfortability was assessed with a single question “How do you manage on the income you have available?” (1 = It is impossible, 2 = It is difficult all the time, 3 = It is difficult some of the time, 4 = It is not too bad, 5 = It is easy).

Infant and birthing details were collected at T3, including: *Postpartum complications* “Did you have any postnatal complications? (Yes/No)”, *Infant’s temperament* “How would you rate your baby’s temperament (10-point scale)” and *Infant’s impact on maternal sleep* “To what extent did the infant have a negative impact on your sleep quality or quantity? (1 = Very little, 2 = A little, 3 = Somewhat, 4 = Much, 5 = Very much)”.

#### Maternal Relationship Satisfaction

Relationship satisfaction was measured using the Dyadic Adjustment Scale 4 item (DAS-4). The DAS-4 is a shortened version of the Dyadic Adjustment Scale consisting of four items, scored on a Likert scale. Items 1-3 ranged from 0 (all the time) to 5 (never), while item 4 ranged from 0 (extremely unhappy) to 6 (perfect). Scores range from 0-21, with higher scores indicative of better relationship satisfaction (Sabourin et al., 2005). DAS-4 scores 13 and below indicates relationship distress.

#### Depression symptoms

Depression symptoms were measured using the Patient-Reported Outcome Measurement Information System Depression Short Form (PROMIS Depression SF). The PROMIS Depression - SF measures emotional distress and depressed mood via eight questions, scored on a 5-point Likert scale (1 = never, 5 = always). Raw scores (range 8-40) are converted to standard *t*-scores ranging from 37.1 to 81.1, with higher scores indicating more depressive symptoms (Pilkonis et al., 2011).

#### Anxiety Symptoms

Anxiety symptoms were measured via the Patient-Reported Outcome Measurement Information System Anxiety Short Form (PROMIS Anxiety – SF; (Pilkonis et al., 2011). Aspects of worry, fear, heightened arousal and associated physiological arousal were assessed with eight questions scored on a 5-point Likert scale (1 = never, 5 = always). Raw scores (range 8-40) were converted to standard *t*-scores ranging from 37.1 to 81.1, with higher scores indicating more anxiety symptoms.

### Data Analysis

Data were analysed using R 4.3.0 (R Core Team, 2023) and Mplus (Muthén, L.K & Muthén, B.O, 2023) via MplusAutomation (Hallquist & Wiley, 2018). Latent growth modelling (LGM) was used to identify the trajectory of relationship satisfaction change over time and missing data were handled using full information maximum likelihood (FIML). The following models were compared for fit to the data. In all models, time scores were specified as months prior to and since birth, with T1 and T2 being −2.5, −1.5, and T3 to T8 being 1.5, 3, 6, 12, 24. Following visual inspection of the raw data, the final timepoint (T8) was freely estimated in all models to capture a plateauing trend between T7 and T8. To evaluate model fit, multiple indices of model fit and relevant suggested cut-offs were considered including Comparative Fit Index (CFI) > 0.95, Root Mean Square Error of Approximation (RMSEA) ≤ 0.06 and Standardised Root Mean Square Residual (SRMR) ≤ 0.08 (Hu & Bentler, 1999). The models were as follows:

*Model 1:* One-piece linear LGM with random intercept and slope.

*Model 2:* Two-piece LGM with one intercept and two slopes: Slope 1 for pregnancy (from T1 to T2) and Slope 2 for postpartum (from T3 to T8).

Model 2 showed significantly better fit to the data (Δχ^2^ = 48.10, *df* = 4, *p* = < 0.0001) compared to Model 1. As model fit was suboptimal, modification indices were consulted and a suggested correlation residual between DAS at T5 and DAS at T6 was added to the model (Model 3), which significantly improved model fit, compared to Model 2 (Δχ^2^ = 13.01, *df* = 1, *p* < 0.0001).

In order to explore factors associated with the trajectory of relationship satisfaction, a further LGM model (Model 4) was constructed with predictors of intercept and slopes added to Model 3. For the following analysis, due to insufficient number of respondents in categories other than Caucasian, ethnicity was dichotomised to race. The predictors included for slope 1, slope 2 and intercept were: race (White as “0” and non-White as “1”), education level (Bachelor degree and below as “0” and postgraduate as “1”), mental health history (no current/past mental health condition as “0” and current or past mental health condition as “1”), financial comfortability (It is impossible/It is difficult all the time/ It is difficult some of the time as “0” and “It is not too bad/It’s easy” as “1”), Intervention allocation (Cognitive Behavioural Therapy for Insomnia as “0” and Healthy Diet Program as “1”). For slope two (postpartum) only predictors included: postpartum complications, infant’s temperament, infant’s impact on maternal sleep (“Very little/A little/Somewhat” as “0” and “Much/Very much” as “1”).

To maximise statistical power and support parsimony, non-significant predictors that failed to demonstrate meaningful associations with intercept or either slope were dropped from subsequent models. The full original model (Model 4) with all predictors is included in Supplementary Material. Based on the above-described steps, the final reported model (Model 5) was conducted using Model 3, with predictors of age, education, financial comfortability and race on the intercept and both slopes.

Finally, to test whether attrition at T8 impacted model predictions, a final supplementary model was conducted as Model 3 but without final timepoint (T8). The results of this model were not different from the model with T8 (Supplementary Material) and thus, Model 3 was retained to allow for inclusion of most available data.

## RESULTS

A total of 157 participants were in a relationship at baseline and thus were included in analysis. All participants were female, were on average 33.36 (*SD* = 3.40) years old, and were predominately white (86.6%). Baseline depression and anxiety scores were below the population mean, indicating low symptom burden (Table 1). There was a drop in participants by T8, where only n = 75 participants reported relationship satisfaction. There was no significant difference in baseline characteristics (age, race, education, financial comfortability, relationship satisfaction) between those who completed the study and those who did not (Supplementary Material). There were small non-significant correlations between baseline age, race, education and financial comfortability (Supplementary Material).

**Table 1.**
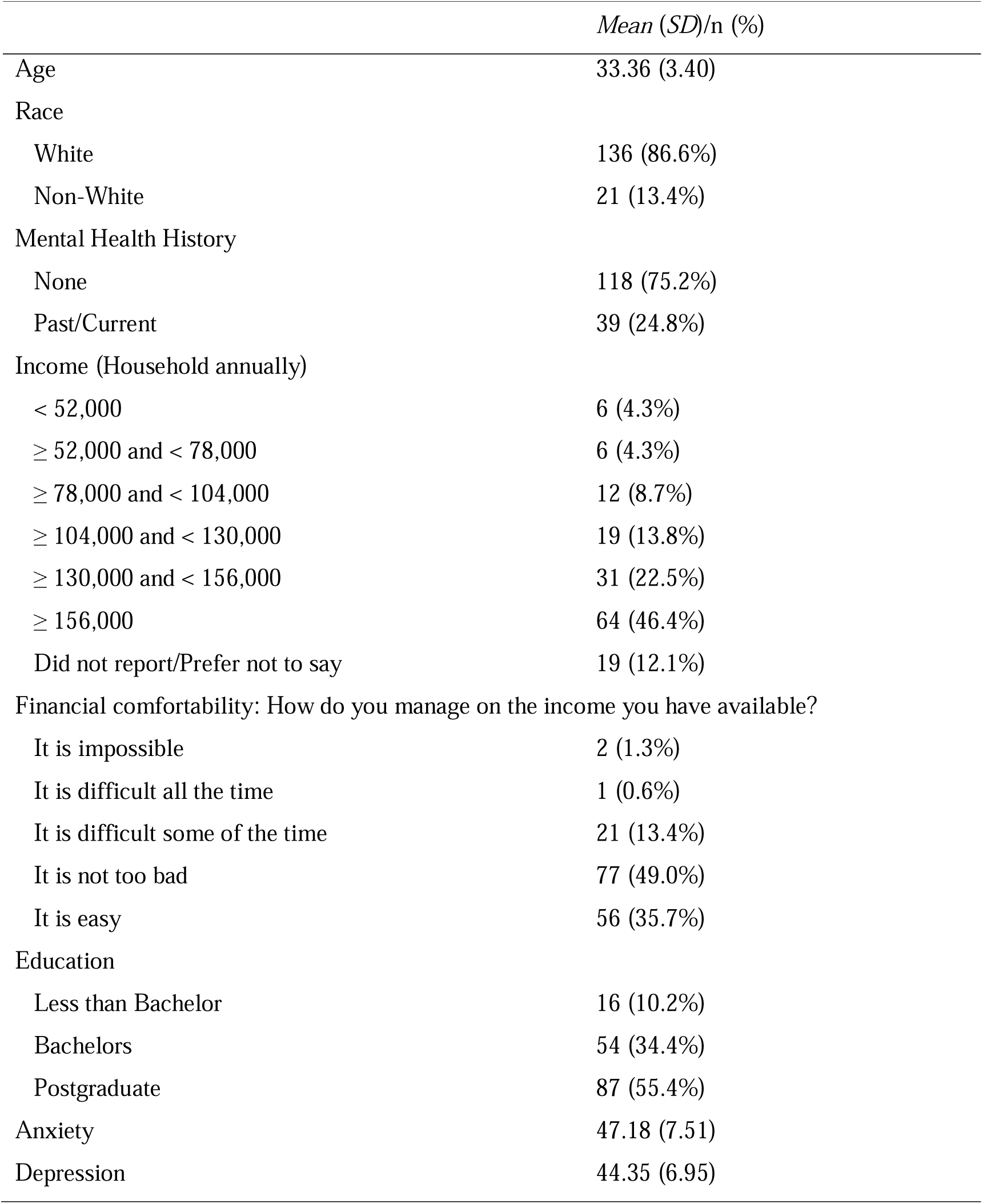
Baseline characteristics of included participants.

### Relationship satisfaction trajectory

Relationship satisfaction at baseline was high 18.44 (1.97). Average relationship satisfaction remained above the clinical distress range (> 13) at every time point (Figure 1). The proportion of participants below the clinically distressed range of relationship satisfaction (≤ 13) increased from 2.5% at baseline to 18.7% at five years postpartum (Table 2).

**Figure 1.**
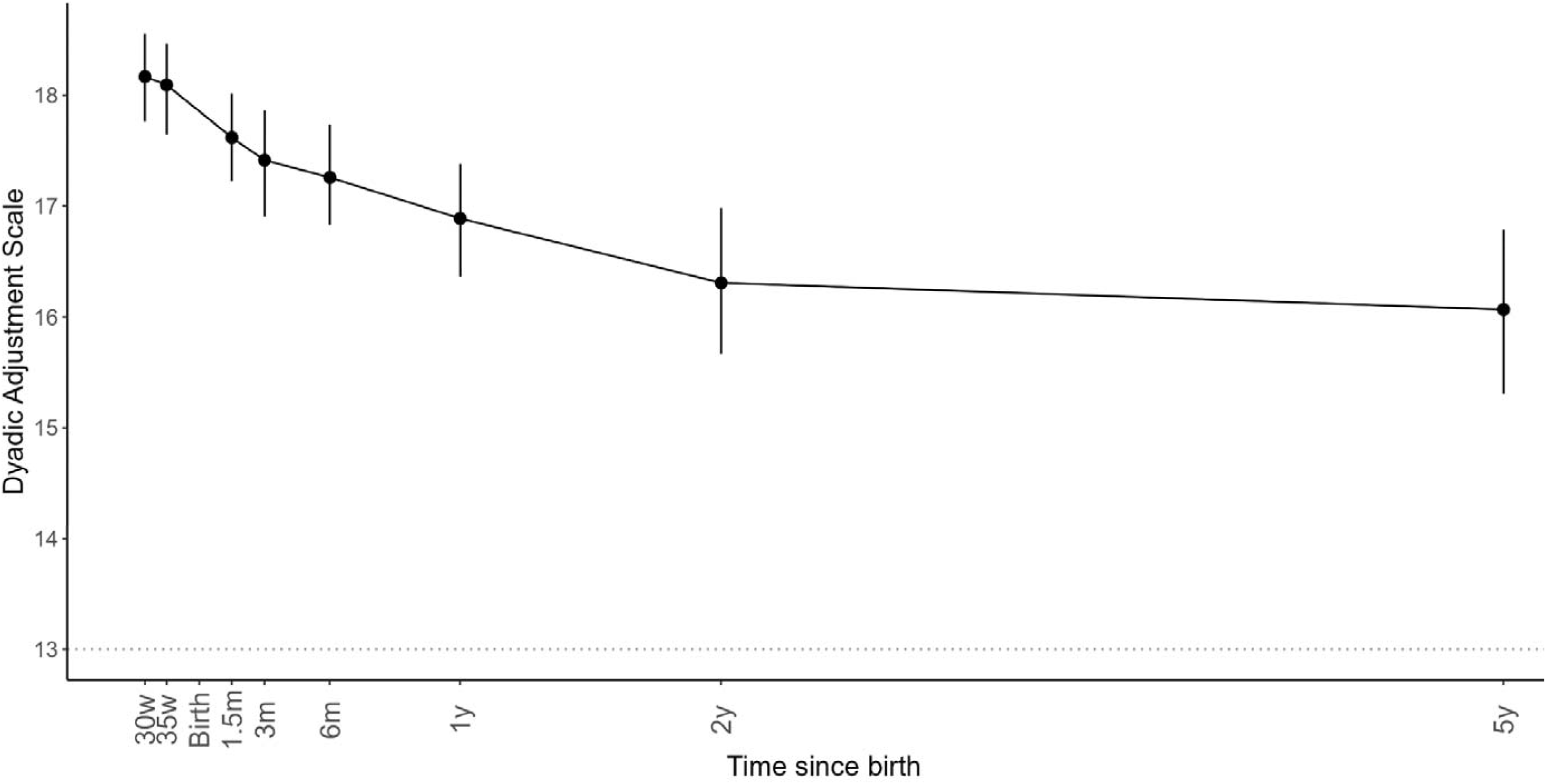
Average Relationship Satisfaction scores of the study period (Dot represents *Mean* and Error Bars are 95% Confidence Intervals). Dotted line denotes “Relationship Distress” point.

**Table 2.** Average relationship satisfaction and percentage of participants in the clinical distress range at each timepoint.

|  | T1<br>(N<br>157) | = T2<br>(n = 148) | T3<br>(n = 149) | T4<br>(n = 145) | T5<br>(n = 145) | T6<br>(n = 119) | T7<br>(n = 109) | T8<br>(n = 75) |
| --- | --- | --- | --- | --- | --- | --- | --- | --- |
| DAS |  |  |  |  |  |  |  |  |
| *Mean* | 18.44 | 18.27 | 17.77 | 17.43 | 17.35 | 16.94 | 16.53 | 16.07 |
| *(SD)* | (1.97) | (2.10) | (2.45) | (2.94) | (2.72) | (2.73) | (3.42) | (3.39) |
| DAS | 4 | 4 | 11 | 11 | 10 | 12 | 11 | 14 |
| $\leq 13$ | (2.5%) | (2.7%) | (7.4%) | (7.6%) | (6.9%) | (10.1%) | (10.1%) | (18.7%) |

Note. DAS = Dyadic Adjustment Scale, $\text{DAS} \leq 13$ indicates clinical distress range. T1 = 30 weeks gestation, T2 = 35 weeks gestation, T3 = 1.5 months postpartum, T4 = 3 months postpartum, T5 = 6 months postpartum, T6 = 12 months postpartum, T7 = 24 months postpartum, T8 = 5 years postpartum.

The final reported models (Model 3, Model 5) had adequate-good fit with CFI >0.95 and RMSEA ≤0.06. However, SRMR was slightly above suggested ideal cut off in the current analysis (M3 = 0.10, M5 = 0.09) (Table 3).

**Table 3.** Model fit statistics for all models.

| Model | X<sup>2</sup>(df), p | RMSEA [90% CI] | CFI | SRMR |
| --- | --- | --- | --- | --- |
| M1 | 98.60 (30) = 0.00 | 0.12 [0.10, 0.15] | 0.88 | 0.28 |
| M2 | 50.51 (26) = 0.00 | 0.08 [0.05, 0.11] | 0.96 | 0.14 |
| **M3** | **37.49 (25) = 0.05** | **0.06 [0.00, 0.09]** | **0.98** | **0.10** |
| M4 | 110.28 (79) = 0.01 | 0.05 [0.03, 0.07] | 0.95 | 0.10 |
| **M5** | **75.37 (48) = 0.01** | **0.06 [0.03, 0.09]** | **0.96** | **0.09** |

Note. All models freely estimated time score at T8. M1: One-piece LGM with random intercept and slope, M2: Two-piece LGM with one intercept and two slopes: Slope 1 for pregnancy (T1 to T2) and Slope 2 for postpartum (T3 to T8). M3: Model 2 with one correlated residual between relationship satisfaction at T5 and T6. M4: Model 3 with all predictors (see Supplementary Material), M5: Final predictor model, M3 + significant predictors from M4 (race, financial comfortability, education, age). **Bold text** denotes models presented in the main manuscript.

Mean relationship satisfaction at birth was estimated to be 17.67 [*95% Confidence Interval* = 17.28-18.06] (Table 4). Both slope 1 and slope 2 were significantly smaller than 0, indicating a significant decline in relationship satisfaction during both late pregnancy prior to birth, and across the postpartum period (Table 4). During late pregnancy, the decline appeared to be at a greater rate at 0.32 point per month, compared to a slower decline at 0.06 point per month during the postpartum follow up (Table 4). The freely estimated time score at T8 was 17.91 [11.20, 24.62], indicating relationship satisfaction at this time (60 months postpartum) was estimated to be comparable to ∼18 months postpartum.

**Table 4.** Unstandardised change trajectories in relationship satisfaction from third trimester to five years postpartum.

| Outcome | Predictor | Estimate [95% CI], *p* |
| --- | --- | --- |
| Mean | Intercept | **17.67 [17.28, 18.06] <0.0001** |
|  | Slope 1 | **-0.32 [-0.46, -0.19] <0.0001** |
|  | Slope 2 | **-0.06 [-0.08, -0.03] <0.0001** |
| Variances | Intercept | **4.80 [3.36, 6.25] <0.0001** |
|  | Slope 1 | 0.19 [-0.01, 0.38] 0.06 |
|  | Slope 2 | **0.01 [0.01, 0.02] 0.001** |
| Slope 1 with Intercept |  | **0.77 [0.35, 1.19] <0.0001** |
| Slope 2 with Intercept |  | -0.05 [-0.11, 0.02] 0.14 |
| Slope 1 with Slope 2 |  | -0.02 [-0.05, 0.00] 0.07 |

Note. Two-piece LGM with one intercept and two slopes: Slope 1 for pregnancy (T1 to T2) and Slope 2 for postpartum (T3 to T8). In this model T8 was freely estimated. **Bold text** denotes significant outcome.

The variance of the intercept was significant (*p* < 0.0001), suggesting large individual differences in overall levels of relationship satisfaction at baseline. Furthermore, the variance of slope 2 was significant (*p* = 0.001), indicating significant individual variations in trajectories across the postpartum period; however, the variance of slope 1 was not significant (*p* = 0.06). There was a significant positive association between the intercept and slope 1 (*p* < 0.0001), indicating that those who started with higher baseline levels experienced less decline. There was no significant association between slope 2 and the intercept or slope 1 with slope 2, suggesting that the rate of relationship satisfaction decline during pregnancy and postpartum were not related to each other.

### Factors associated with relationship satisfaction trajectory

As shown in Table 5, being of non-White (vs White) race was associated with overall lower relationship satisfaction (*p* = 0.004) by approximately 1.6 points. In the postpartum (slope 2), several factors were associated steeper decline in relationship satisfaction (Table 5). Greater financial comfort (*p* = 0.007) and holding a postgraduate degree (*p* = 0.049) predicted a slower decline in relationships satisfaction over time. Conversely, older maternal age at baseline predicted a significantly steeper decline in postpartum relationship satisfaction (*p* = 0.026).

**Table 5.** Unstandardised trajectory of relationship satisfaction with baseline predictors.

| Outcome | Predictor | Estimate [95% CI], *p* |
| --- | --- | --- |
| Intercept | **Race** | **-1.61 [-2.70, -0.52] 0.004** |
|  | Financial Comfortability | 0.64 [-0.44, 1.72] 0.25 |
|  | Education Level | -0.38 [-1.13, 0.37] 0.32 |
|  | Age | -0.01 [-0.12, 0.10] 0.87 |
| Slope 1 | Race | -0.11 [-0.41, 0.20] 0.49 |
|  | Financial Comfortability | -0.19 [-0.48, 0.10] 0.20 |
|  | Education Level | 0.03 [-0.17, 0.23] 0.78 |
|  | Age | 0.03 [0.00, 0.06] 0.07 |
| Slope 2 | Race | 0.05 [0.00, 0.10] 0.06 |
|  | **Financial Comfortability** | **0.07 [0.02, 0.11] 0.007** |
|  | **Education Level** | **0.03 [0.00, 0.07] 0.049** |
|  | **Age** | **-0.01 [-0.01, 0.00] 0.026** |

Note. Two-piece LGM with one intercept and two slopes: Slope 1 for pregnancy (T1 to T2) and Slope 2 for postpartum (T3 to T8) with correlated residuals for relationship satisfaction over time, with predictors: Race (White = 0, Non-white = 1), Financial comfortability (Less comfortable = 0, More comfortable = 1), Education level (Less than postgraduate = 0, Postgraduate = 1), Age at baseline. **Bold text** denotes significant outcome.

## DISCUSSION

The present study examined the trajectory of relationship satisfaction in first time mothers from late pregnancy to five years postpartum. There was a significant decrease in relationship satisfaction from pregnancy to birth and from birth to five years postpartum. By five years postpartum 18.7% of the sample reported within the clinical distress range of relationship satisfaction. Nevertheless, on average, relationship satisfaction did not fall within the level of clinical distress in the present sample. Being non-White was associated with lower overall relationship satisfaction, whilst younger age, higher education, and greater financial comfort were protective against a decline in the postpartum period. It is crucial to note, however, our sample was predominately White, well-educated and of above average income, which limits generalisability to broader samples.

Majority of participants reported high levels of relationship satisfaction at baseline, and despite an overall decline, the average level of relationship satisfaction was not at the clinically distressed level. However, our findings indicate that relationship satisfaction continued to decline, and the number of participants reporting clinical distress levels of relationship satisfaction increased, up to and beyond two years postpartum and remained below baseline and birth levels. This observed decline, based on longitudinal data, is consistent with previous literature based on cross-sectional data that found parents with children in early childhood have lower relationship satisfaction than parents of infants (Han et al., 2023; Meyer et al., 2016). These findings are also consistent with existing studies that suggest for some individuals the transition to parenthood may be associated with substantial relationship decline (Mitnick et al., 2009; Twenge et al., 2003) and for some, marriage breakdown (Helland et al., 2014). Nevertheless, it is worth noting that the decline in relationship satisfaction between two years and five years postpartum was minimal (Supplementary Material) and may have even recovered slightly. This plateau may reflect a stabilisation of relationship satisfaction following the acute decline in the first two years postpartum.

This study identified several factors related to relationship satisfaction trajectories. Being of non-White race was associated with overall poorer relationship satisfaction. Minority populations across the world have been shown to experience poorer maternal outcomes (De Freitas et al., 2020; Kildea et al., 2012). While not examined directly in the current study, it is possible that a lack of support (Coast et al., 2016) or availability of culturally appropriate prenatal practices (Kildea et al., 2012) may have contributed to the relationship satisfaction decline. It is imperative that culturally appropriate prenatal and birthing practices are in place to support parents of minority groups in the transition to parenthood (Coast et al., 2016). This finding must be interpreted within the distinct context of our sample characteristics, which poses a limitation to generalisability. Our cohort was predominantly White (86.6%) and of above average income, which may underestimate systematic stressors and protective factors relevant to marginalised or economically diverse mothers. Therefore, there is a clear need for future studies to utilise more purposeful sampling for more diverse samples, to better understand this relationship.

In this study, older parents, those less financially comfortable and those with less than postgraduate education experienced a steeper decrease in relationship satisfaction. These results align with previous literature suggesting that socioeconomic factors moderate relationship satisfaction during times of stress (Maisel & Karney, 2012). Financial pressures have been cited as a challenge during the transition to parenthood (Lévesque et al., 2020) and uniquely predict relationship dissolution during early childhood parenting (Helland et al., 2014). It is important to note that our sample was highly educated, with 89.8% obtaining a bachelor’s degree or higher, which is higher than the rate of 45-51% in Australian women aged between 24-44 years old (Australian Bureau of Statistics, 2021). As such these findings may not generalise to other samples. Although all participants were experiencing the transition to parenthood for the first time, older parents reported a slightly steeper relationship satisfaction decline. While some literature suggests that older parents may have had more experience in their relationships with their partners, enabling them to better cope with the challenges of parenthood (Doss et al., 2009), older parents in our sample may be experiencing unique pressures. For instance, older parents may have spent longer periods accustomed to established routines, making the demands of welcoming a first child more disruptive. Interestingly, there was no significant correlation between age, education, race and financial comfortability in the current sample, indicating that these may play an important role independent of each other.

Overall, our exploratory analysis of baseline and perinatal factors provides valuable preliminary insights into factors that may be associated with relationship satisfaction during this period. Predictors of the intercept of relationship satisfaction, such as race, identify baseline disparities during late pregnancy, highlighting which couples may have existing vulnerabilities during pregnancy. Conversely, associates of the slope/s, such as education, financial comfortability and age, represent factors which may play a role as drivers or buffers to relationship satisfaction during this time.

Importantly, many of the original psychosocial and physiological predictors included in the baseline predictor model (e.g., postpartum complications, infant’s temperament, infant’s impact on maternal sleep; see Supplementary Material) did not show a significant relationship with change trajectory. While these findings suggest that these factors did not emerge as drivers within our specific sample, they do not preclude the potential clinical relevance of infant and maternal stressors. The lack of statistical significance for these factors, may reflect unique sample characteristics or lack of statistical power to detect subtle effects in the broader model, rather than the absence of a relationship. There is a need for further research which explores predictors of relationship satisfaction change during this period.

Finally, while there was an overall significant decline in relationship satisfaction, the change for most was minimal and for majority of our sample, they remained well above the level of “distress” (range of those above “clinically distress” threshold at all timepoints 81.3%-97.5%). Furthermore, our final timepoint estimate (five years) showed that while relationship satisfaction did not return to mean baseline levels, it had returned to a level approximately equal to 18 months postpartum. As such, our findings highlight that while relationship satisfaction may decline and may not return to baseline levels, the change was small within our current sample.

### Limitations

The findings of the current study must be interpreted in light of some limitations. First and foremost, the sample is largely White, of high education and socioeconomic status, and further research is needed for samples of diverse background. There is a clear need for more diverse samples to better understand changes in relationship satisfaction across many different cultural and contextual environments for mothers.

The current study also did not examine relationship satisfaction from a dyadic perspective, which would have provided unique insights into the dynamic interactions of a relationships over time during an important life stage. The high level of relationship satisfaction from baseline, while reflective of many couples during pregnancy period (Tissera et al., 2021), means that the findings from this study are likely not generalisable to those experiencing relationship distress prior to welcoming a child.

Additionally, while our study is strengthened by including five years postpartum, the lapse of time between the two year and five year follow ups means that we may have missed nuanced changes in relationship satisfaction during this time. Future studies may benefit from more regular data collection after two years postpartum. Furthermore, although missing data were handled using FIML, attrition at five year time point was large. Sensitivity analyses showed that findings are consistent when analyses are carried to two year postpartum, suggesting that attrition at the final time point is not likely to have meaningfully affected overall findings.

It is worth noting that while our primary model fit indices (CFI and RMSEA) indicated acceptable fit, the SRMR was slightly elevated. As SRMR is sensitive to sample size (Pavlov et al., 2021), given our modest sample size, this elevated SRMR likely reflects an artifact of small sample size rather than poor model fit.

Finally, our analysis did not consider whether parents had a subsequent child during the follow up period. It is possible that some parents within the sample did welcome a subsequent baby during this time, which may have had additional impacts on relationship satisfaction (Twenge et al., 2003).

Future explorations should consider subsequent pregnancy, birth and timing of pregnancy and birth and potential impacts on relationship satisfaction across this period.

## Conclusion

The current study demonstrates that relationship satisfaction declines from pregnancy to five years postpartum in a sample of Australian mothers. Minority race, financial comfortability, education level and age may influence the magnitude of this trajectory. Despite overall high relationship satisfaction, nearly one in five of the sample showed clinical relationship distress at five years postpartum. These findings highlight a need to better support couples during this challenging time, not just through pregnancy and birth, but also through early childhood.

## Data Availability

Data are not available publicly due to ethics restrictions.

## ACKNOWLEDGEMENTS

Authors would like to thank every mother and infant for generously donating their time, and the following individuals in their contribution to the larger project (alphabetical): Laura Astbury, Michelle Blumfield, Kaye Dyson, Cassandra Fong, Catherine Fulgoni, Elisabeth Gasparini, Anthony Hand, Ashley Lam, Jin Joo Lee, Wing (Theodora) Leung, Rachel Manber, Sarah Meiklejohn, Allie Peters, Monika Raniti, Sarah Samuel, Lin Shen, Isabelle Smith, Emma Thompson, Sumedha Verma, Giovanna Wolten, Addie Wootten.

## ETHICS DECLARATIONS

### Ethics Approval

This study was conducted in line with the Australian National Statement on Ethical Conduct in Human Research (2007). Ethics approval was attained from the Royal Women’s Hospital (#16/01) and Monash University Human Research Ethics Committees (#CF16/1561– 2016000815).

### Consent to participate

All participants provided informed consent for themselves and their infant to participate, prior to enrolment in study.

### Consent for publication

Informed consent to participate in study included consent for publication of de-identified results.

### Clinical Trial Number

This study is retrospectively registered (ACTRN12616001462471) on 19/10/2016, after the first participant enrolled on 16/5/2016 but before the end of recruitment on 11/01/2017. The delay in registration was due to limited resources during the early phase of recruitment, when recruitment rate was substantially higher than expected.

### Author Contribution

M.E.C: Conceptualization, Formal analysis, Methodology, Writing - original draft, and Writing - review & editing. S.C.O.W.S: Conceptualization, and Writing - review & editing. D.M.P: Conceptualization, Investigation, Project administration, and Writing - review & editing. S.P.A.D: Conceptualization, Investigation, and Writing - review & editing. B.B: Conceptualization, Investigation, Funding acquisition, Methodology, Project administration, Formal analysis, Supervision, and Writing - review & editing.

