## Supplementary Material for "TRAJECTORY AND PREDICTORS OF MATERNAL RELATIONSHIP SATISFACTION FROM PREGNANCY TO FIVE YEARS POSTPARTUM"

Supplementary Table 1. Results of Latent Growth Model 4, all predictors included in model

| Outcome | Predictor | Estimate [*95% CI*], *p* |
| --- | --- | --- |
| Intercept | Intervention Group | -0.23 [-0.96, 0.50] 0.54 |
|  | **Race** | **-1.67 [-2.74, -0.60] 0.002** |
|  | Financial Comfortability | 0.11 [-0.97, 1.19] 0.85 |
|  | Education Level | -0.27 [-1.01, 0.48] 0.49 |
|  | Age | -0.01 [-0.11, 0.10] 0.89 |
|  | Mental Health History | -0.69 [-1.49, 0.12] 0.09 |
| Slope 1 | Intervention Group | -0.14 [-0.33, 0.05] 0.14 |
|  | Race | -0.06 [-0.35, 0.23] 0.70 |
|  | Financial Comfortability | -0.10 [-0.37, 0.18] 0.51 |
|  | Education Level | 0.01 [-0.18, 0.20] 0.91 |
|  | Age | 0.03 [0.00, 0.05] 0.07 |
|  | Mental Health History | -0.08 [-0.29, 0.13] 0.44 |
| Slope 2 | Intervention Group | 0.02 [-0.01, 0.05] 0.16 |
|  | **Race** | **0.05 [0.00, 0.10] 0.045** |
|  | **Financial Comfortability** | **0.06 [0.02, 0.11] 0.008** |
|  | **Education Level** | **0.03 [0.00, 0.07] 0.036** |
|  | **Age** | **0.00 [-0.01, 0.00] 0.033** |
|  | Mental Health History | 0.02 [-0.02, 0.05] 0.34 |
|  | Infant Sleep Impact | 0.01 [-0.02, 0.04] 0.42 |
|  | Postpartum Complications | 0.00 [-0.03, 0.02] 0.78 |
|  | Infant Temperament | -0.02 [-0.05, 0.01] 0.12 |

Supplementary Table 2. Results of Latent Growth Model 6, supplementary analysis of only timepoints T1-T7

| Outcome | Predictor | Estimate [*95% CI*], *p* |
| --- | --- | --- |
| **Mean** | **Intercept** | **17.67 [17.28, 18.06] <0.0001** |
|  | **Slope 1** | **-0.32 [-0.46, -0.19] <0.0001** |
|  | **Slope 2** | **-0.05 [-0.08, -0.03] <0.0001** |
| **Variances** | **Intercept** | **4.81 [3.36, 6.26] <0.0001** |
|  | Slope 1 | 0.19 [0.00, 0.38] 0.05 |
|  | **Slope 2** | **0.01 [0.01, 0.02] <0.0001** |
| **Slope 1 with Intercept** | | **0.77 [0.36, 1.19] <0.0001** |
| Slope 2 with Intercept | | -0.06 [-0.13, 0.01] 0.08 |
| **Slope 1 with Slope 2** | | **-0.03 [-0.05, 0.00] 0.047** |

Supplementary Table 3. Comparison of baseline characteristics between those who completed final timepoint and those who did not.

|  | Completed (n=75) | Did not complete  (n=82) | *p* |
| --- | --- | --- | --- |
| Baseline DAS | 18.61 (1.97) | 18.53 (1.58) | 0.81 |
| Age | 33.72 (4.06) | 33.04 (2.80) | 0.25 |
| Race |  |  | 0 .84 |
| White | 59 (89.4%) | 66 (90.4%) |  |
| Non-white | 7 (10.6%) | 7 (9.6%) |  |
| Financial comfortability | |  | 0.81 |
| More comfortable | 57 (86.4%) | 62 (84.9%) |  |
| Less comfortable | 9 (13.6%) | 11 (15.1%) |  |
| Education |  |  | 0.23 |
| Postgraduate | 41 (62.1%) | 38 (52.1%) |  |
| Less than postgraduate | 25 (37.9%) | 35 (47.9%) |  |

Supplementary Table 4. Pearson’s R correlations and p-values between predictors in final model

|  | Age | Education | Financial comfortability | Race |
| --- | --- | --- | --- | --- |
| Age | 1.00 | 0.17 (*p* = 0.05) | 0.08 (*p* = 0.36) | -0.03 (*p =* 0.76) |
| Education |  | 1.00 | 0.06 (*p* = 0.51) | 0.00 (*p* = 0.98) |
| Financial  comfortability | | | 1.00 | 0.14 (*p* = 0.12) |
| Race |  |  |  | 1.00 |
